# European-derived coronary artery disease polygenic scores over-flag genetic risk in Vietnamese and Southeast Asian populations: a multi-score analysis in 1000 Genomes

**DOI:** 10.64898/2026.07.10.26357796

**Authors:** Quý Hoàng Phú, Thân Lê Xuân, Dũng Ðoàn Ðúc

## Abstract

**Background:** Polygenic scores (PRS) for coronary artery disease (CAD) are derived almost entirely from European-ancestry data. Their portability to Southeast Asian populations, including the Vietnamese, is largely uncharacterised and clinically consequential when scores are used with risk thresholds.

**Methods:** We evaluated four independent European-derived CAD scores from the PGS Catalog (PGS000058, PGS000349, PGS002809, PGS004198; 70–5,723 variants) in 2,504 individuals from the 1000 Genomes Project, focusing on the Vietnamese Kinh (KHV) and Dai (CDX) samples. Per-individual scores were computed with PLINK2 and standardised. We assessed (i) the cross-ancestry distribution (calibration) and (ii) a clinically-relevant consequence: the proportion of each population flagged “high genetic risk” when the European top-20% threshold is applied (20% if perfectly calibrated).

**Results:** For the primary score (PGS000058) the standardised PRS differed across super-populations (ANOVA F(4, 2499) = 121.1, p < 0.001); the Vietnamese Kinh mean was +0.47 SD above the European mean (Welch t = 7.77, p = 2.0×10^−14^). Applying the European top-20% high-risk threshold, the fraction of Vietnamese Kinh flagged ranged from 22.2% to 57.6% across the four scores, and of Dai from 21.5% to 43.0%, versus the intended 20%. Three of the four scores over-flagged Vietnamese (25–58%); the largest score (PGS004198) was approximately calibrated for East/Southeast Asians (∼22%) but markedly over-flagged Africans (69.3%).

**Conclusions:** European-derived CAD polygenic scores are inconsistently calibrated in Vietnamese and other Southeast Asian samples, and most substantially over-flag high genetic risk when a European threshold is applied. The magnitude and even the direction of miscalibration depend on the specific score, so no such score can be assumed transferable without local validation and recalibration. Distribution shift bounds, but does not by itself quantify, loss of predictive accuracy, which requires phenotyped data.

## 1. Introduction

Coronary artery disease is a leading cause of death worldwide and has a substantial heritable component. Polygenic scores aggregate the effects of many variants into a single estimate of inherited risk and, when combined with a threshold, are increasingly proposed for clinical risk stratification. However, the discovery datasets behind most CAD scores are overwhelmingly of European ancestry.

Polygenic scores are known to lose accuracy and calibration across ancestries because allele frequencies, linkage-disequilibrium structure and sometimes effect sizes differ between populations. Southeast Asian populations, including the more than 97 million people of Vietnam, are among the most under-represented in genomic reference resources. If existing CAD scores are miscalibrated in these groups, threshold-based clinical use could systematically over- or under-identify individuals as high risk and widen health disparities.

Using only openly available data—genotypes from the 1000 Genomes Project and four published scores from the PGS Catalog—we evaluated how European-derived CAD polygenic scores behave in the Vietnamese Kinh (KHV) and Dai (CDX) samples. Because the 1000 Genomes samples have no cardiovascular phenotype, we restrict the analysis to calibration and to the *risk-stratification consequence* of that calibration (the fraction flagged by a European threshold), and specify the phenotyped-cohort analysis needed to quantify predictive accuracy.

## 2. Materials and Methods

### 2.1 Data sources

All analyses used publicly available, de-identified data; no new human data were collected. Data sources are summarised in Table 1.

**Table 1.** Data sources.

| Resource | Content used | Access |
| --- | --- | --- |
| 1000 Genomes phase 3 (GRCh37) | Genotypes for 2,504 individuals incl. KHV (Vietnamese Kinh, n=99) and CDX (Dai, n=93) | Open download |
| PGS Catalog | Four European-derived CAD scores: PGS000058, PGS000349, PGS002809, PGS004198 (harmonized scoring files) | Open, no application |
| GenomeAsia 100K / national biobank | Individual-level genotypes + CAD phenotype (for the discrimination extension only) | Controlled (EGA) |

### 2.2 Genotype data and population definitions

Genotypes were taken from the 1000 Genomes Project phase 3 call set (GRCh37; 2,504 individuals), grouped by the project’s population and super-population labels. The Southeast-Asia-relevant target populations were KHV (Kinh in Ho Chi Minh City, Vietnam; n = 99) and CDX (Chinese Dai in Xishuangbanna; n = 93); EUR, EAS, SAS, AMR and AFR served as comparators.

### 2.3 Polygenic scores and computation

Four independent European-derived CAD scores were retrieved from the PGS Catalog via its REST API using harmonized GRCh37 scoring files: PGS000058 (204 genome-wide-significant variants; Morieri et al. 2018), PGS000349 (70 variants; Pechlivanis et al. 2020), PGS002809 (205 variants; Ahmed et al. 2022) and PGS004198 (5,723 variants, LASSO-derived; Raben et al. 2023). Per-individual scores were computed in PLINK2 by allele-matched dosage weighting. The numbers of variants successfully matched to the genotype data were 201, 68, 203 and 5,638 respectively (≥97% of each score).

### 2.4 Statistical analysis

For each score, raw scores were standardised to mean zero and unit variance across the pooled sample. We summarised the standardised score by population and super-population and, for the primary score, tested for differences across super-populations by one-way ANOVA. To capture the clinical consequence of miscalibration, we defined a “high genetic risk” threshold as the 80th percentile of the European distribution (the common “top-20%” cut-point) and computed the percentage of each population exceeding it; under perfect cross-ancestry calibration this would be 20% in every group. Analyses were performed in R; all code is openly available (Section 6).

#### Scope of inference

Because the 1000 Genomes samples lack cardiovascular outcomes, we did not estimate discrimination (AUC) or effect size (odds ratio per SD). Section 4 specifies the phenotyped-cohort design required for those estimates.

## 3. Results

### 3.1 Distribution of the primary CAD score across ancestries

For PGS000058, the standardised CAD PRS differed significantly across super-populations (one-way ANOVA F(4, 2499) = 121.1, p < 0.001; Table 2, Figure 1). The score was shifted upward in East and South Asian samples and downward in African and admixed American samples relative to Europeans. The Vietnamese Kinh (KHV) mean was +0.47 SD and the Dai (CDX) mean +0.43 SD above the European mean; at the super-population level East Asians exceeded Europeans by +0.45 SD (Welch t = 7.77, df = 955, p = 2.0×10□^1^□; 95% CI 0.34–0.57). Dispersion was similar across groups, so the difference is a shift in location rather than spread.

**Table 2.**
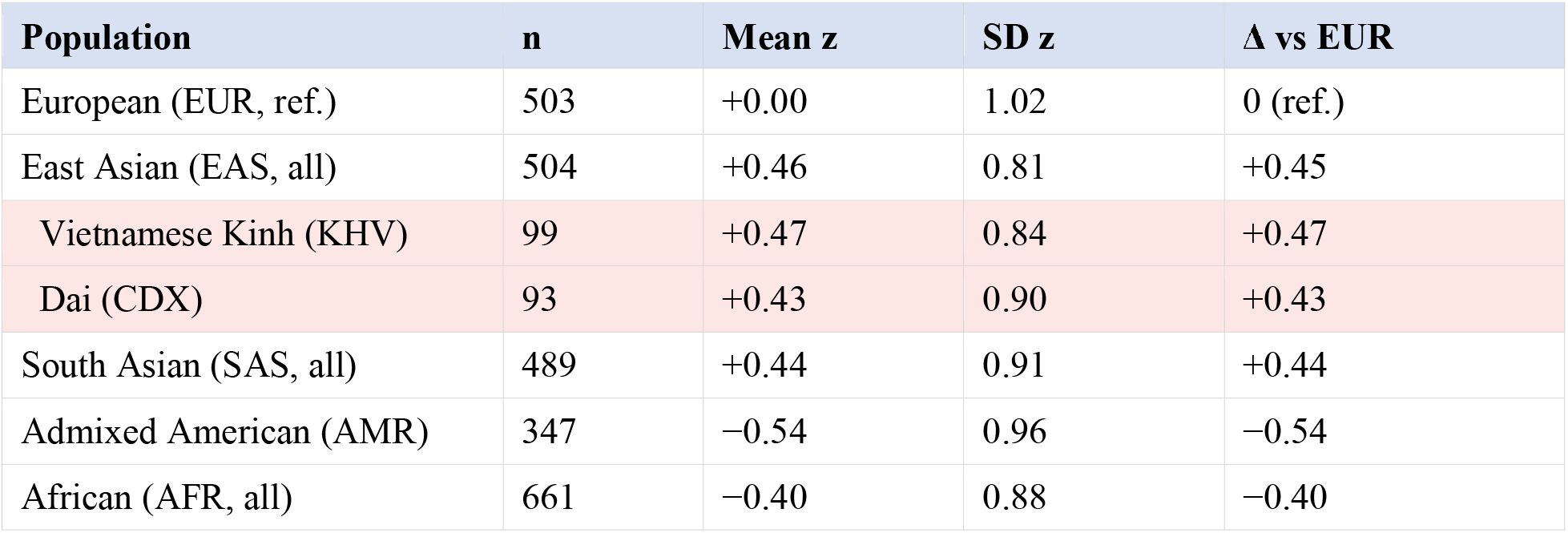
Standardised CAD polygenic score (PGS000058) by population. Southeast Asian target populations shaded.

| Population | n | Mean z | SD z | $\Delta$ vs EUR |
| --- | --- | --- | --- | --- |
| European (EUR, ref.) | 503 | +0.00 | 1.02 | 0 (ref.) |
| East Asian (EAS, all) | 504 | +0.46 | 0.81 | +0.45 |
| Vietnamese Kinh (KHV) | 99 | +0.47 | 0.84 | +0.47 |
| Dai (CDX) | 93 | +0.43 | 0.90 | +0.43 |
| South Asian (SAS, all) | 489 | +0.44 | 0.91 | +0.44 |
| Admixed American (AMR) | 347 | −0.54 | 0.96 | −0.54 |
| African (AFR, all) | 661 | −0.40 | 0.88 | −0.40 |

**Figure 1.**
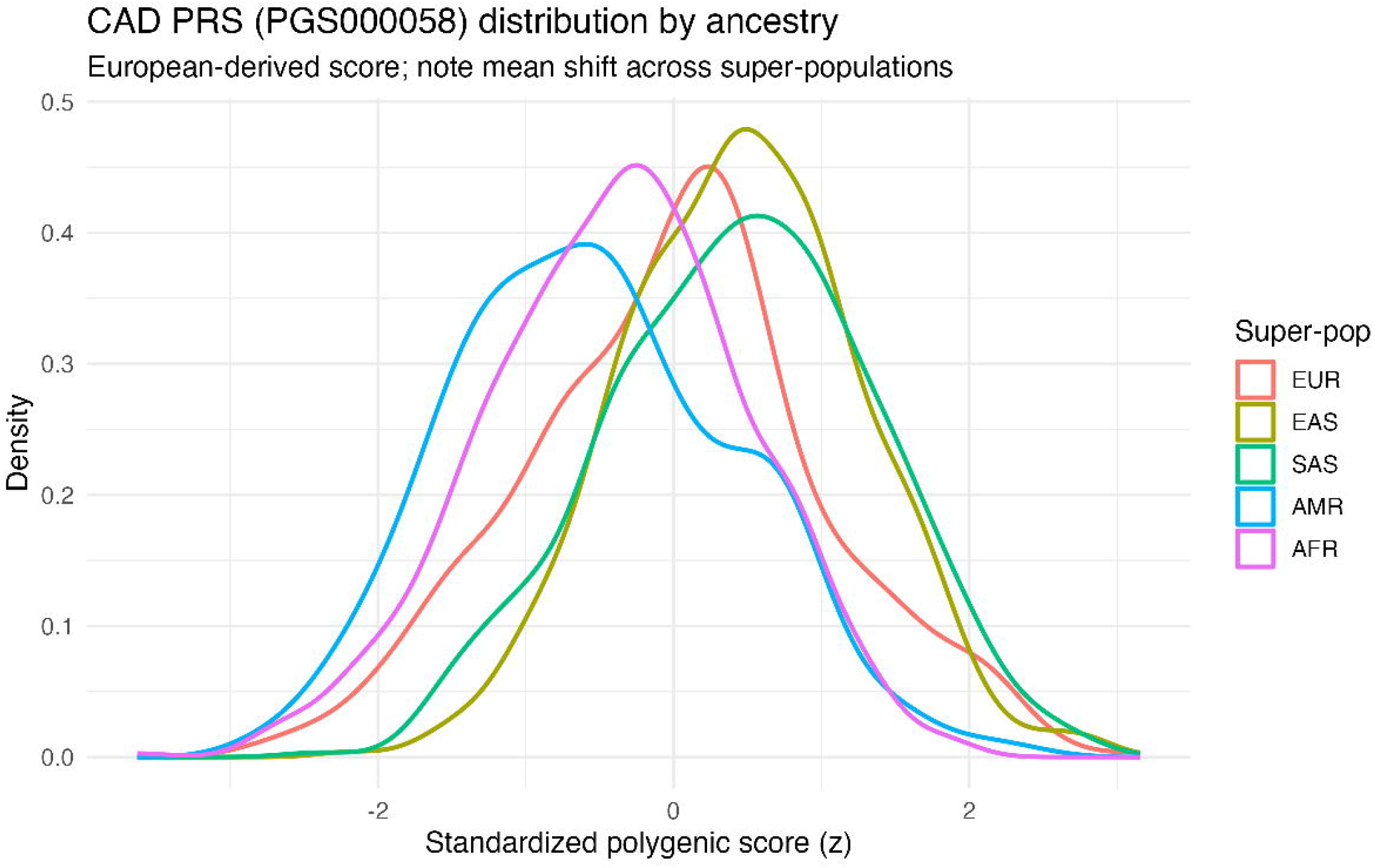
Kernel-density distribution of the standardised CAD polygenic score (PGS000058) by super-population; the European-derived score is shifted upward in East Asian (containing the Vietnamese Kinh and Dai) and South Asian samples.

**Figure 2.**
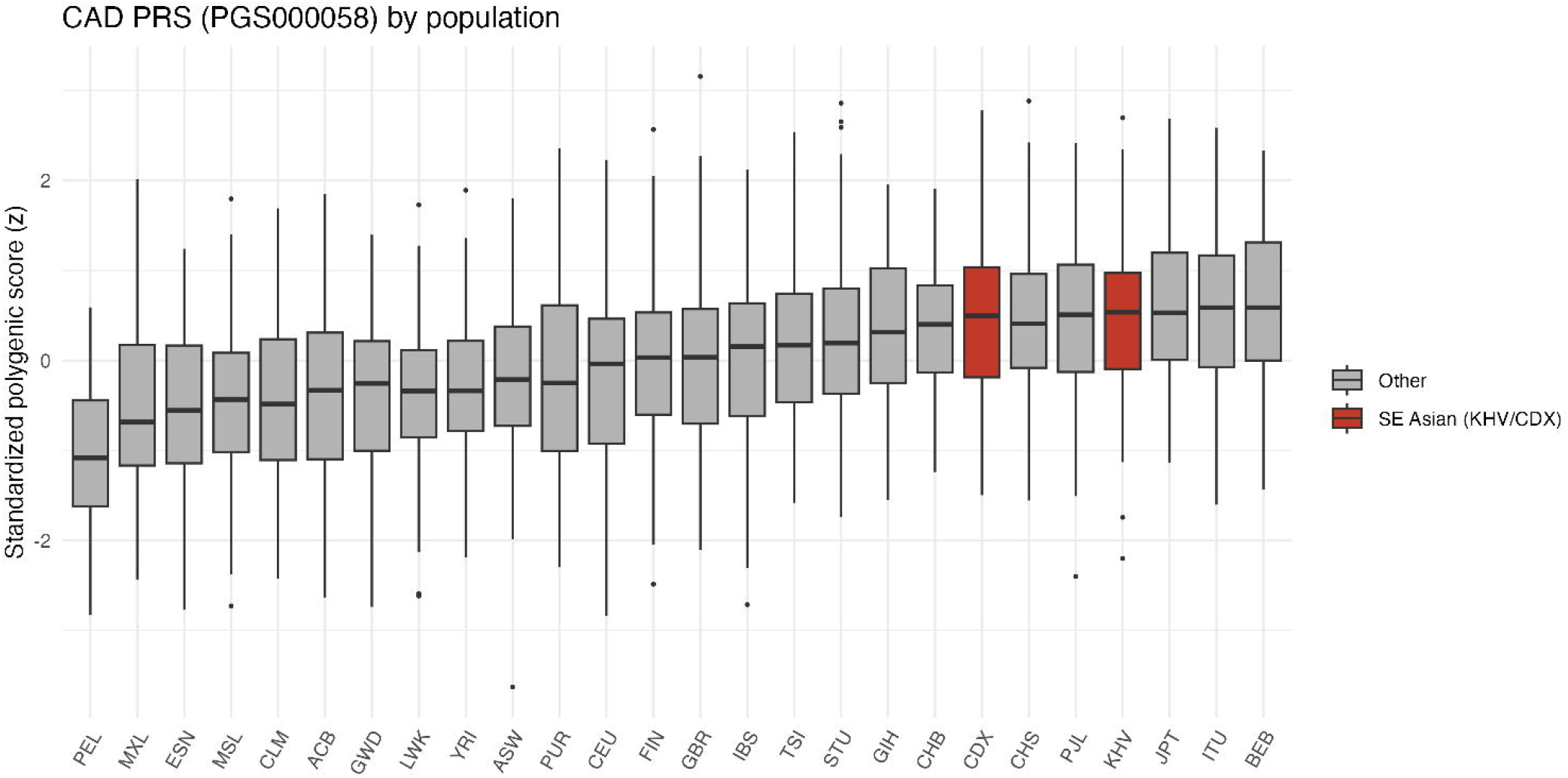
Standardised CAD polygenic score (PGS000058) by 1000 Genomes population, ordered by mean; the Vietnamese Kinh (KHV) and Dai (CDX) samples are highlighted and sit above the European populations.

### 3.2 Consistency across four independent scores and clinical mis-stratification

We repeated the analysis for four independent European-derived CAD scores and quantified the clinical consequence using the European top-20% high-risk threshold, at which a well-calibrated score flags 20% of every ancestry (Table 3, Figure 3). Calibration in the Southeast Asian samples was inconsistent and predominantly toward over-flagging. The percentage of Vietnamese Kinh flagged as high genetic risk ranged from 22.2% to 57.6% across the four scores, and of Dai from 21.5% to 43.0%, versus the intended 20%. Three of the four scores over-flagged Vietnamese: PGS000349 (a 70-variant score) flagged 57.6% of Kinh and 57.0% of Dai—almost three times the intended rate—while PGS000058 flagged 35.4% and 43.0%. The largest score, PGS004198 (5,723 variants, LASSO), was approximately calibrated for East/Southeast Asians (KHV 22.2%, CDX 21.5%) yet dramatically over-flagged Africans (69.3%).

**Table 3.** Percentage of each ancestry flagged “high genetic risk” using the European top-20% threshold (calibrated = 20%). KHV = Vietnamese Kinh, CDX = Dai.

| Score (variants) | EUR | EAS | KHV | CDX | SAS | AFR |
| --- | --- | --- | --- | --- | --- | --- |
| PGS000058 (204) | 20.1 | 34.7 | 35.4 | 43.0 | 36.4 | 9.1 |
| PGS000349 (70) | 20.1 | 59.3 | 57.6 | 57.0 | 38.0 | 49.2 |
| PGS002809 (205) | 20.1 | 23.0 | 25.3 | 28.0 | 19.6 | 5.1 |
| PGS004198 (5,723) | 20.1 | 19.0 | 22.2 | 21.5 | 22.5 | 69.3 |

**Figure 3.**
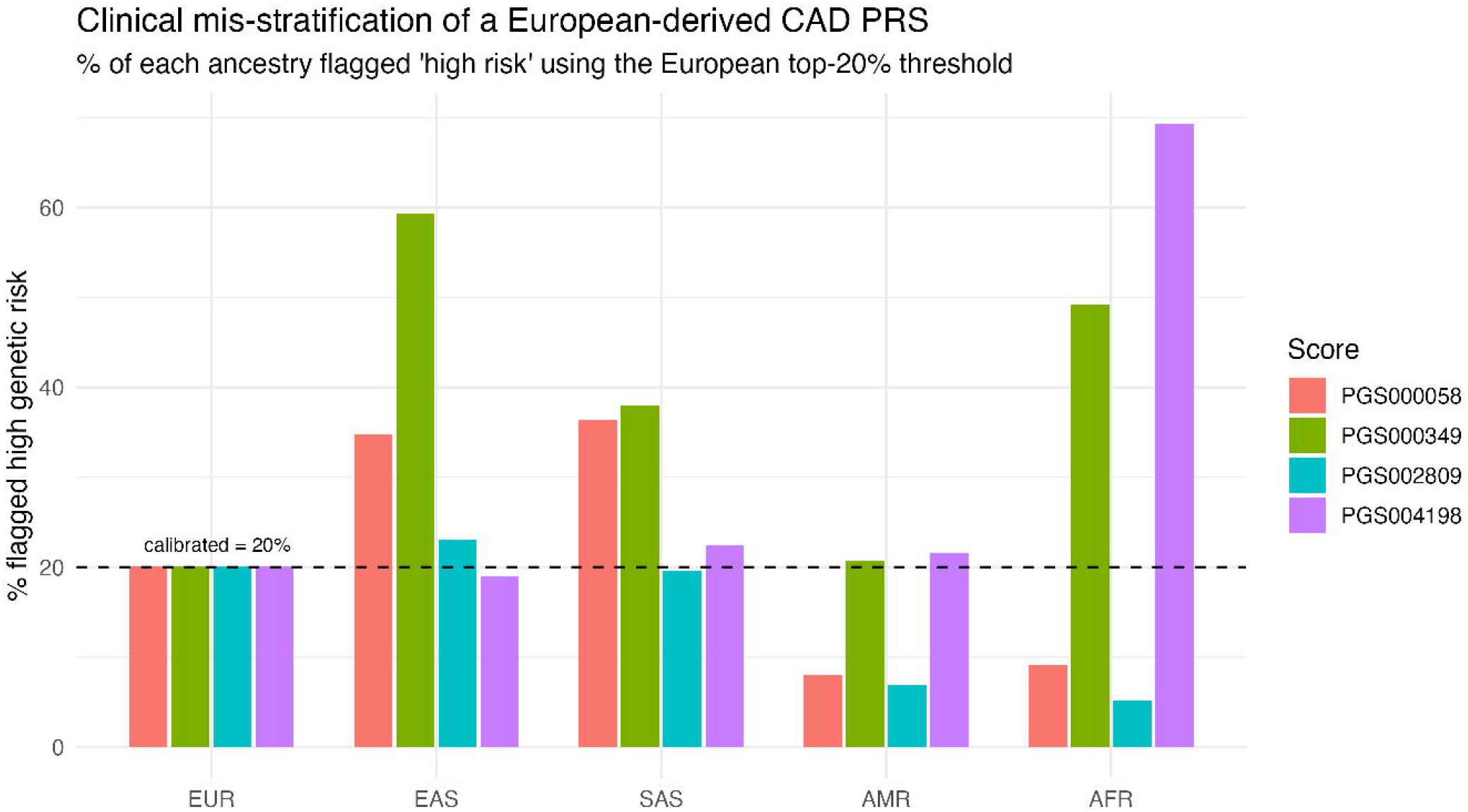
Percentage of each ancestry flagged as high genetic risk when the European top-20% CAD-PRS threshold is applied, for four independent scores. The dashed line marks the calibrated expectation (20%). Bars above the line indicate over-flagging.

## 4. Discussion

Across four independent European-derived CAD polygenic scores, calibration in Vietnamese and other Southeast Asian samples was inconsistent and mostly biased toward over-flagging. For the primary score the distribution was shifted upward by roughly half a standard deviation in Vietnamese Kinh and Dai samples, and applying a European high-risk threshold flagged a substantial excess of these individuals— up to nearly three times the intended rate for one score. Because the scores were built and calibrated in Europeans, this excess reflects allele-frequency and linkage-disequilibrium differences rather than demonstrated higher true risk.

Two features stand out. First, the phenomenon is not an artefact of a single score: three of four independent scores over-flagged Vietnamese, strengthening the conclusion. Second, the magnitude and even the direction of miscalibration depended heavily on the score—one large LASSO-based score was approximately calibrated for East/Southeast Asians but severely over-flagged Africans—so cross-ancestry behaviour cannot be predicted from a score’s European performance and must be checked per score and per population.

### Interpreting distribution versus accuracy

A shifted distribution and threshold mis-stratification demonstrate miscalibration but do not by themselves measure predictive accuracy. Quantifying accuracy requires a Southeast Asian cohort with genotypes and CAD status, in which one can estimate the odds ratio per standard deviation and the area under the ROC curve, stratified by ancestry and adjusted for age, sex and genetic principal components. Such data are available through controlled-access resources (e.g. GenomeAsia 100K via the European Genome-phenome Archive, or national biobanks); the companion code contains the ready-to-run model.

### Implications

Threshold-based CAD polygenic scores calibrated in Europeans should not be deployed in Vietnamese or Southeast Asian care without local recalibration and validation, as most would over-identify high genetic risk and could drive over-referral and over-treatment. The results reinforce the need for larger Southeast Asian genomic and phenotypic resources and for ancestry-specific score development.

#### 4.1 Limitations

First, the 1000 Genomes samples are modest in size and lack cardiovascular phenotypes, so we assess calibration and threshold mis-stratification rather than predictive accuracy. Second, KHV and CDX are single reference samples and may not capture the full diversity of Vietnam or mainland Southeast Asia. Third, we examined four scores spanning 70–5,723 variants; very large genome-wide scores may behave differently and warrant confirmatory work. Fourth, ≥97% of each score’s variants were available, so missingness is unlikely to explain the findings.

## 5. Conclusion

European-derived coronary artery disease polygenic scores are inconsistently calibrated in Vietnamese and Dai samples and, for most scores examined, substantially over-flag high genetic risk when a European threshold is applied. Ancestry-aware recalibration, per-score validation, and phenotyped Southeast Asian cohorts are needed before such scores can be used equitably in the region.

## Data Availability

All data analysed are publicly available. Genotype data are from the 1000 Genomes Project (https://www.internationalgenome.org/); the coronary artery disease polygenic scores (PGS000058, PGS000349, PGS002809, PGS004198) are from the PGS Catalog (https://www.pgscatalog.org/). The complete analysis code and derived results are archived on Zenodo (DOI: 10.5281/zenodo.21304620).

https://doi.org/10.5281/zenodo.21304620

https://www.internationalgenome.org/

https://www.pgscatalog.org/

## 6. Data and code availability

All data are public: the 1000 Genomes Project (internationalgenome.org) and the PGS Catalog (pgscatalog.org; scores PGS000058, PGS000349, PGS002809, PGS004198). The complete analysis pipeline (download, scoring, statistics and figures) that reproduces every result and figure is archived on Zenodo (DOI: 10.5281/zenodo.21304620) and runs end to end on a laptop.

## 7. Declarations

### Ethics approval and consent to participate

Not applicable. This study is a secondary analysis of publicly available, fully de-identified data from the 1000 Genomes Project and the PGS Catalog. No new participants were recruited; informed consent and ethical approvals were obtained by the original studies at the point of data collection, and no additional institutional review board approval was required for this secondary analysis.

### Consent for publication

Not applicable.

### Trial registration

Not applicable. This work is not a clinical trial, so no trial registration was required.

### Availability of data and materials

All data analysed are publicly available: the 1000 Genomes Project (internationalgenome.org) and PGS Catalog scores PGS000058, PGS000349, PGS002809 and PGS004198 (pgscatalog.org). The full analysis pipeline reproducing every result and figure is archived on Zenodo (DOI: 10.5281/zenodo.21304620).

### Authors’ contributions

Q.H.P. conceived and designed the study, assembled the data, performed the analyses, produced the figures and drafted the manuscript. T.L.X. and D.Ð.D. contributed to the interpretation of the results and critically revised the manuscript for important intellectual content. All authors read and approved the final version.

## Acknowledgements

The authors thank the 1000 Genomes Project and the PGS Catalog for making their data openly available.

